# Validation of Machine-Learning Angiography-Derived Physiological Pattern of Coronary Artery Disease

**DOI:** 10.1101/2024.10.17.24315610

**Authors:** Yueyun Zhu, Simone Fezzi, Norma Bargary, Daixin Ding, Roberto Scarsini, Mattia Lunardi, Concetta Mammone, Max Wagener, Angela Mcinerney, Gabor Toth, Gabriele Pesarini, David Connolly, Flavio Ribichini, Shengxian Tu, William Wijns, Andrew J Simpkin

**Author notes:** Equally contributed as joint first authors (Yueyun Zhu, Simone Fezzi). ML-based physiological patterns of disease. Corresponding authors: Prof. William Wijns, The Lambe Institute for Translational Research, Curam and Saolta University Healthcare Group, Galway National University of Ireland Galway (NUIG), Costello Road, Shantalla, Galway, H91 V4AY.; Prof. Andrew J Simpkin, School of Mathematical and Statistical Sciences, Centre for Data Analytics, National University of Ireland, Galway, Ireland, 3.

## Abstract

**Background:** The classification of physiological patterns of coronary artery disease (CAD) is crucial for clinical decision-making, significantly affecting the planning and success of percutaneous coronary interventions (PCI).

**Objectives:** This study aimed to develop a novel index to reliably interpret and classify physiological CAD patterns based on virtual pullbacks from single-view Murray’s law-based quantitative flow ratio (μFR) analysis.

**Methods:** The pullback pressure gradient index (PPGi) was used to classify CAD patterns, with a cut-off value of PPGi=0.78 distinguishing focal from diffuse and non-focal disease. A machine learning method using penalized logistic regression models was proposed to assess CAD patterns. Scores derived from multivariate functional principal component analysis (MFPCA) of μFR and quantitative coronary analysis improved model performance. Expert panel interpretations served as the reference.

**Results:** A total of 179 vessels (134 patients) underwent classification. The PPGi cut-off of 0.78 achieved 70% accuracy (95% CI: 0.70 to 0.71) for focal vs. diffuse and 77% accuracy (95% CI: 0.76 to 0.77) for focal vs. non-focal classification. The penalized logistic regression model, including PPGi as a feature, provided superior accuracy: 95% (95% CI: 0.94 to 0.95) for focal vs. diffuse and 84% (95% CI: 0.83 to 0.84) for focal vs. non-focal classification. Positive predictive value (PPV) and negative predictive value (NPV) were 95% and 92% (focal vs. diffuse) and 84% each (focal vs. non-focal). Overall, the penalized logistic regression model successfully identified more focal lesions and ensured fewer diffuse or non-focal lesions were misclassified.

**Conclusions:** The machine learning method with penalized logistic regression outperformed the PPGi cut-off values, providing robust and generalizable classification across different study populations.

## Introduction

The classification of physiological patterns of coronary artery disease (CAD), including focal, serial lesions, diffuse disease and mixed patterns, has become increasingly important in clinical decision making (1–5). Recent studies have investigated the role of coronary physiological indices in assessing CAD patterns, such as wire-based fractional flow reserve (FFR), instantaneous wave-free ratio (iwFR) or quantitative flow ratio (QFR) computed from the coronary angiogram (1–5). Qualitative interpretation of physiological pattern is influenced by the expertise of physicians, and subject to inter- and intra-operator variability [8]. Such difficulties in standardizing CAD patterns definitions and classification, led to the development of different mathematical metrics to classify physiological patterns, including FFR gradient per unit time (dFFR[t]/dt) and pullback pressure gradient index (PPGi) (4, 6–11). These metrics were applied to determine the disease severity along the course of the vessel and predict post-PCI outcomes. Both these metrics are calculated based on pressure-wire (PW)-pullback performed during continuous hyperaemia. PPGi quantitatively measures the physiological distribution of coronary plaques along the vessel and is capable of distinguishing between focal and diffuse disease, while dFFR[t]/dt reflects the local physiological disease severity. Despite the extensive evidence in support of physiological assessment as a gate-keeper for coronary revascularization, and the growing one in support of physiology guidance for percutaneous coronary intervention (PCI) optimization, its use remains hampered by economic and logistic reasons (i.e., need for a PW and for hyperaemic agents, increased costs and procedural time) (12–14). Notably, both the PPGi and dFFR[t]/dt metrics require automatized motorized PW-pullback, further limiting the applicability in the real-world practice.

To overcome such limitations, angiography-derived physiological indices, such as the Murray’s law-based quantitative flow ratio (μFR), have been recently developed and validated (3, 14, 15). μFR provides an inherently co-localized virtual pullback that qualitatively interpret the physiological pattern of disease (focal vs. diffuse vs. mixed vs. serial) (2, 3, 16, 17). A lack of consensus in the qualitative interpretation of physiological pattern definition is evident, making it challenging to reach a standardization, relying on operator interpretation and experience.

The aim of this study was to propose a novel machine learning method to quantify and classify physiological patterns, which can be reproducible and generalized across a range of study samples, having qualitative interpretation as a reference standard.

## Methods

### Study population

This is a proof-of-concept, validation study including a cohort of patients with native coronary atherosclerosis that underwent clinically-driven percutaneous coronary intervention (PCI) between December 2012 and September 2017 at the Verona University Hospital. Patients younger than 18 years old, with known contraindication to dual anti-platelet therapy, a concomitant indication to open-heart surgery, heart transplanted patients with allograft vasculopathy or patients presenting with resuscitated cardiac arrest and women with childbearing potential were excluded from the study. The inclusion criteria for PCI were defined as the presence of at least one critical (degree of stenosis; DS≥70%) or intermediate de novo lesion (DS between 50% and 70%) detected at the basal coronary angiography associated with concomitant clinical signs or symptoms, or any intermediate lesion resulted hemodynamically relevant (≤0.80) as assessed by FFR.

The study was conducted in accordance with the ethical principles of the Declaration of Helsinki, and it was approved by the institutional ethical board of the Verona University Hospital (185 CESC). All the patients had provided their written consent for the anonymous data collection.

Overall, the study utilized coronary angiographic data from 134 patients (179 vessels) included in previously reported studies (18). For each patient, information such as age, gender, smoking status, hypertension status and diabetes status were provided.

### Murray law-based quantitative flow ratio analysis

Coronary angiograms were anonymized, sent, and centrally analyzed in the Smart Sensors Lab (Lambe Institute for Translational Medicine, University of Galway). Computation of μFR was performed using the μFR software (AngioPlus Core, version V3, Pulse Medical, Shanghai, China) by an experienced analyst, blinded to any clinical data. Angiographic exclusion criteria were ostial disease in the left main or in the right coronary artery, target vessel with collateral circulation or coronary flow from patent surgical grafts, target vessel with myocardial bridging, target vessel with previous myocardial infarction and poor angiography image quality. The detailed methodology for single-view μFR computation has been described previously (19). Routine cut-off value of μFR (≤0.80) was used to classify the hemodynamic significance. For each vessel, fundamental descriptors such as lesion length, diameter and %DS at the quantitative coronary analysis (QCA) were included.

### Physiological pattern of CAD interpretation

Physiological patterns of CAD indexes were derived from μFR virtual pullback traces: physiological distribution was assessed through the QFR-virtual pullback PPGi which discriminates predominantly focal from predominantly diffuse disease, providing a continuous metric based on the magnitude of maximum pressure drop over 20 mm and on the extent of functional disease over the entire interrogated vessel.

PPGi was calculated as follows:

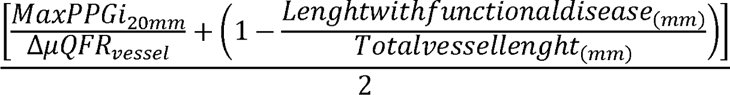

As previously reported (16), high PPGi values (close to 1) suggest predominantly focal disease, whereas low values (close to 0) predominantly diffuse disease. PPGi cut-off value (0.78) was used to dichotomize CAD into focal (PPGi ≥ 0.78) and diffuse (PPGi <0.78) disease, as previously validated (20, 21).

Local physiological severity was calculated by the instantaneous μFR gradient per unit length (dμFR/ds), using a cutoff value of 0.025/mm to identify the presence (≥0.025/mm) or the absence (<0.025/mm) of major gradients, as previously validated (20, 21).

In order to develop a novel machine learning-based computational tool for the interpretation of physiological pattern of CAD, μFR values were extracted from the pressure tracing every 0.35 mm. Together with the μFR values along the vessels’ length, a point-by-point (every 0.35 mm) reconstruction of further quantitative coronary analysis (QCA)-derived variables was obtained (i.e., minimal lumen diameter, reference vessel diameter). Vessels with distal μFR>0.95 were considered without functional disease and excluded from the current analysis.

### Qualitative Physiological patterns interpretation

In this study, 179 vessels were assessed independently by eight expert cardiologists, with significant experience in acquisition and interpretation of coronary physiology. Using a dedicated offline portal, experts interpreted each trace independently, without access to the coronary angiogram or additional clinical information, resulting in a qualitative interpretation of physiological pressure drop over the vessel, using previously proposed definitions as a reference (22). Each expert labelled each vessel as having one of four distinct physiological CAD patterns, namely focal lesion, diffuse disease, mixed pattern and serial lesions. The final decision for each physiological pattern was determined by the interpretation provided from the eight cardiologists, defined as the agreement of at least 5 out of 8, see **Figure 1**. At the first round assessment, there were 63 vessels (34%) with a consensus reached by all eight cardiologists, and 153 vessels (83%) with an agreement from at least 5 cardiologists. The remaining 32 vessels (17%) with less than 5/8 concordant interpretations underwent a second stage of assessment until the divergences between the cardiologists were solved. The determinations made by the eight expert cardiologists were regarded as the reference standard, which was utilized to train the machine learning algorithm and provide the classification results obtained in later sections (23).

**Figure 1.**
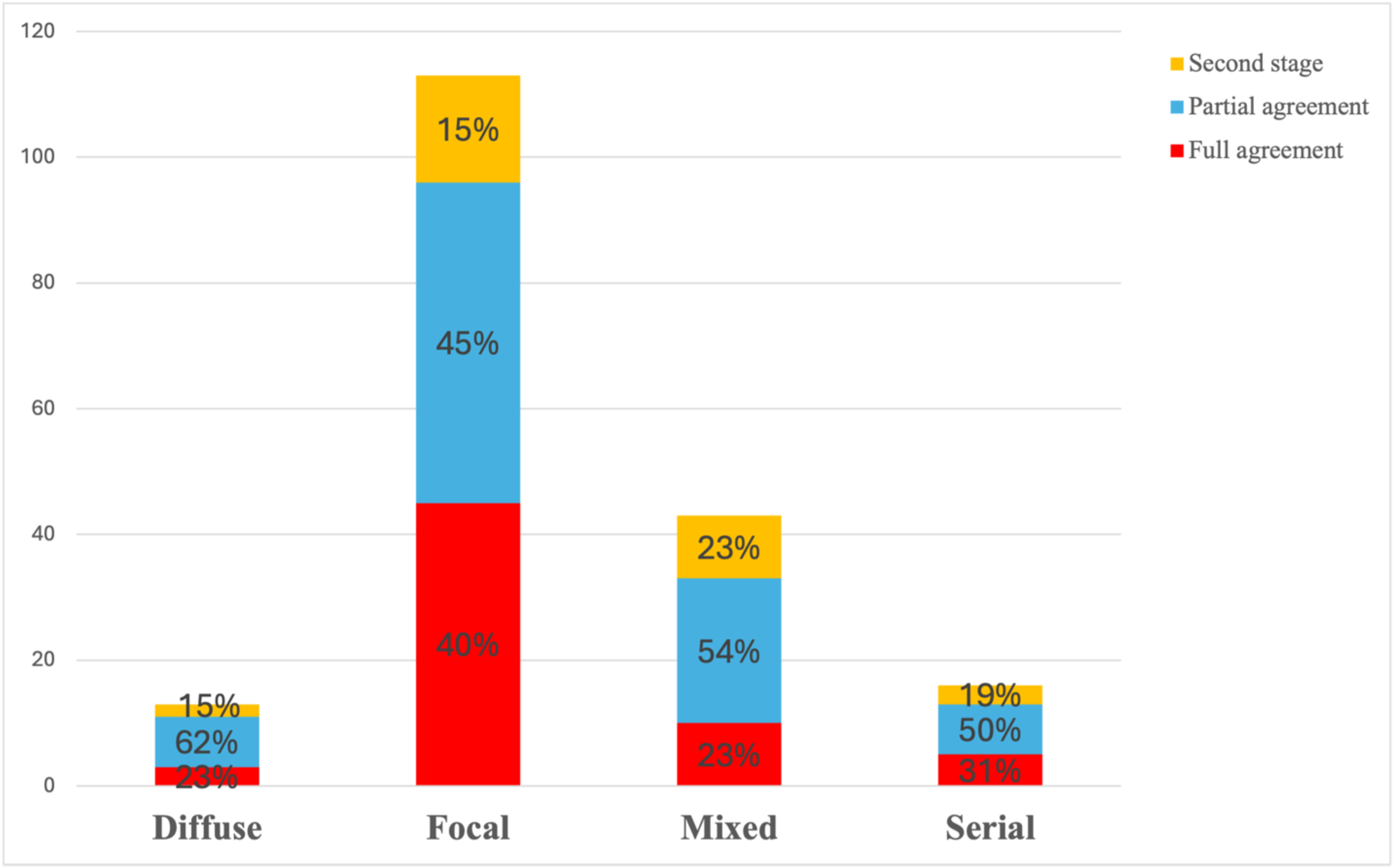
Qualitative interpretation of coronary artery disease patterns obtained from eight experienced interventional cardiologists. At the first round assessment, full agreement was considered a consensus reached by all eight cardiologists (red), while partial as the agreement from at least 5 cardiologists. Vessels with less than 5/8 concordant interpretations underwent a second stage of assessment until the divergences between the cardiologists were solved. The determinations made by the eight expert cardiologists were regarded as the reference standard, which was utilized to train the machine learning algorithm and provide the classification results obtained in later sections.

### Study endpoints

Overall, the study aimed to develop a machine learning method for physiological pattern interpretation based on a single-view angiography-derived physiological assessment, in order to attain high-level classification performance and reproducibility.

The first physiological pattern of CAD classification concentrated on differentiating focal from diffuse disease. The second classification dealt with focal vs non-focal patterns (including diffuse, mixed and serial lesions). Due to the imbalanced number among four physiological patterns (see **Table 1**), fitting a multinomial classification caused a lack of accuracy, particularly for the two minority classes: diffuse (n = 12 vessels, 7%) and serial (n = 16 vessels, 9%) patterns. Therefore, it was reasonable to combine diffuse, serial and mixed patterns as a non-focal class and then, perform a binary classification on focal vs non-focal cases.

**Table 1.** Baseline characteristics.

|  | Total<br>(N=179) | Diffuse 12<br>(7%) | Focal 109<br>(61%) | Mixed 42<br>(23%) | Serial 16<br>(9%) |
| --- | --- | --- | --- | --- | --- |
| **Patient characteristics** |  |  |  |  |  |
| Age | 55 ± 11 | 54 ± 8 | 54 ± 12 | 57 ± 11 | 58 ± 12 |
| Sex male | 159<br>(89%) | 11<br>(92%) | 100<br>(92%) | 32<br>(76%) | 16<br>(100%) |
| Non smoker | 59<br>(33%) | 7<br>(58%) | 37<br>(34%) | 9<br>(21%) | 6<br>(38%) |
| Current smoker | 84<br>(47%) | 4<br>(33%) | 49<br>(45%) | 25<br>(60%) | 6<br>(38%) |
| Previous smoker | 36<br>(20%) | 1<br>(8%) | 23<br>(21%) | 8<br>(19%) | 4<br>(25%) |
| Hypertension | 99<br>(55%) | 8<br>(67%) | 50<br>(46%) | 28<br>(67%) | 13<br>(81%) |
| Diabetes | 25<br>(14%) | 3<br>(25%) | 11<br>(10%) | 11<br>(26%) | 0<br>(0%) |
| Dyslipidemia | 115<br>(64%) | 7<br>(58%) | 69<br>(63%) | 29<br>(69%) | 10<br>(62%) |
| **Clinical presentation** |  |  |  |  |  |
| Chronic coronary syndrome | 28<br>(16%) | 1<br>(8%) | 18<br>(17%) | 6<br>(14%) | 3<br>(19%) |
| Unstable angina | 15<br>(8%) | 1<br>(8%) | 10<br>(9%) | 2<br>(5%) | 2<br>(12%) |
| Myocardial infarction | 135<br>(75%) | 10<br>(83%) | 80<br>(73%) | 34<br>(81%) | 11<br>(69%) |
| **Vessel characteristics** |  |  |  |  |  |
| Maximum length | 81.95 ± 23.87 | 82.5 ± 24.44 | 78.12 ± 24.53 | 88.77 ± 20.98 | 89.71 ± 21.98 |
| μFR | 0.68 ± 0.17 | 0.73 ± 0.19 | 0.67 ± 0.17 | 0.69 ± 0.14 | 0.65 ± 0.13 |
| Minimum diameter | 1.25 ± 0.33 | 1.47 ± 0.34 | 1.19 ± 0.32 | 1.35 ± 0.31 | 1.22 ± 0.3 |
| Reference diameter | 3.47 ± 0.68 | 3.49 ± 0.74 | 3.46 ± 0.68 | 3.47 ± 0.54 | 3.58 ± 0.95 |
| PPGi | 0.75 ± 0.12 | 0.59 ± 0.13 | 0.82 ± 0.08 | 0.65 ± 0.09 | 0.67 ± 0.09 |
| dμFR/ds | 0.08 ± 0.06 | 0.03 ± 0.03 | 0.09 ± 0.06 | 0.06 ± 0.05 | 0.08 ± 0.04 |

*Note:* Values are mean ± standard deviation or n (%).
dμFR/ds, instantaneous μFR gradient per unit length; μFR, Murray's law-based quantitative flow ratio; PPGi, pullback pressure gradient index

### Statistical analysis

In this study, two penalized logistic models with elastic net regularization (24) were developed, (see Supplemental Table 1). The first model, namely μFR-model, included thirteen clinical and procedural features. Two features were related to vessel characteristics (reference diameter and maximum length), six features were related to demographical information (age, gender, smoking, diabetes, dyslipidemia and hypertension status), three features were the scores estimated from multivariate functional principal component analysis (MFPCA), and the last two features were μFR and dμFR/ds. The second model, named PPGi-model, extended μFR-model by including an additionally feature PPGi. Detailed statistical methods and models are described in Supplementary Materials.

All analyses were performed in R version 4.3.0 (R Foundation for Statistical Computing). The package MFPCA (25) was used to calculate multivariate functional principal components and scores. The package glmnet (26) was used for the penalized logistic regression model. The overall performance of the models on classification was assessed by accuracy, sensitivity, specificity, positive predictive value (PPV), negative predictive value (NPV), Gwet’s AC1 (27) and area under the curve (AUC) values. The AC1 coefficient introduced by Gwet (27) is more robust when assessing the inter-rater reliability compared to Cohen’s Kappa, particularly for imbalanced data, as AC1 is less affected by prevalence and marginal probability.

## Results

### Baseline characteristics

The baseline characteristics of the study population (134 patients with 179 vessels) were summarized in **Table 1**. The mean age was 55 ± 11 years, and 89% of the patients were men. Based on the experts committee’ s adjudication, 109 vessels (61%) were deemed to have focal CAD patterns and 70 (39%) were identified with non-focal patterns. Within the non-focal patterns, 12 vessels were classified as diffuse, and therefore, a total of 121 vessels were assessed to have either focal or diffuse CAD patterns.

For focal and serial lesions, μFR values were lower than the average for the entire population. PPGi value was significantly lower in diffuse patterns, as compared to focal lesions (0.58 ± 0.12 vs. 0.82 ± 0.08; p value <0.01).

### Classification using PPGi cut-off values

Based on the decisions from eight cardiologists, 121 vessels were adjudicated to have focal or diffuse patterns. The distribution of PPGi across these 121 vessels is shown in **Figure 3a**, which indicates the distribution was negatively skewed due to the imbalanced number of the focal and diffuse patterns. The red dashed line in the left plot of **Figure 3** is the cut-off value of PPGi=0.78. Following this, the 121 vessels were subjected to the classification, of which 74 focal (sensitivity 68%) and 11 diffuse (specificity 92%) were correctly classified, leading to an accuracy of 70% (95% CI: 0.61 to 0.78), with PPV at 99% and NPV at 24%.

The distribution of PPGi across 179 vessels with focal and non-focal patterns is shown in **Figure 3b**, with the red dashed line at value of 0.78. By using PPGi=0.78 cut-off to distinguish focal vs. non-focal disease, 74 focal (sensitivity 68%) and 64 non-focal (specificity 91%) were correctly classified, resulting in an accuracy of 77% (95% CI: 0.70 to 0.83), with PPV at 93% and NPV at 65%.

### Classification using μFR-model

To prevent model from overfitting, 121 vessels with focal and diffuse disease were randomly split into training (n=91, 75%) and testing (n=30, 25%) sets. The μFR-model was fitted to the training data and used to classify the focal and diffuse patterns for the testing set. μFR-model achieved 87% accuracy (95% CI: 0.69 to 0.96), with 25 focal (sensitivity 93%) and 1 diffuse (specificity 33%) correctly classified, see **Figure 4b**. PPV and NPV were 93% and 33%, respectively. The receiver operating characteristic curve (ROC) curve with an AUC of 0.81 (p-value = 0.04) was provided in **Figure 2**. Moreover, the same testing set was assessed by the cut-off value of PPGi=0.78, which correctly classified all 3 diffuse (specificity 100%) and 17 focal (sensitivity 63%), see **Figure 4a**, leading to an accuracy of 67%, PPV of 100% and NPV of 23%. For the classification of focal and non-focal vessels, 179 vessels were randomly partitioned into training (n=135, 75%) and testing (n=44, 25%) sets. **Figure 4a** and **b** illustrate more focal lesions (n=20, sensitivity 74%) were correctly classified from μFR-model compared to the method using PPGi=0.78 as the cut-off value (n=15, sensitivity 56%). In contrast, less non-focal lesions (n=9, specificity 53%) were successfully assessed by μFR-model compared to the method using PPGi=0.78 (n=16, specificity 94%). Overall, the use of μFR-model resulted in a decrease in accuracy, PPV and NPV compared to the cut-off value PPGi=0.78, declining from 70% to 66%, 94% to 71%, 57% to 53%, respectively.

**Figure 2.**
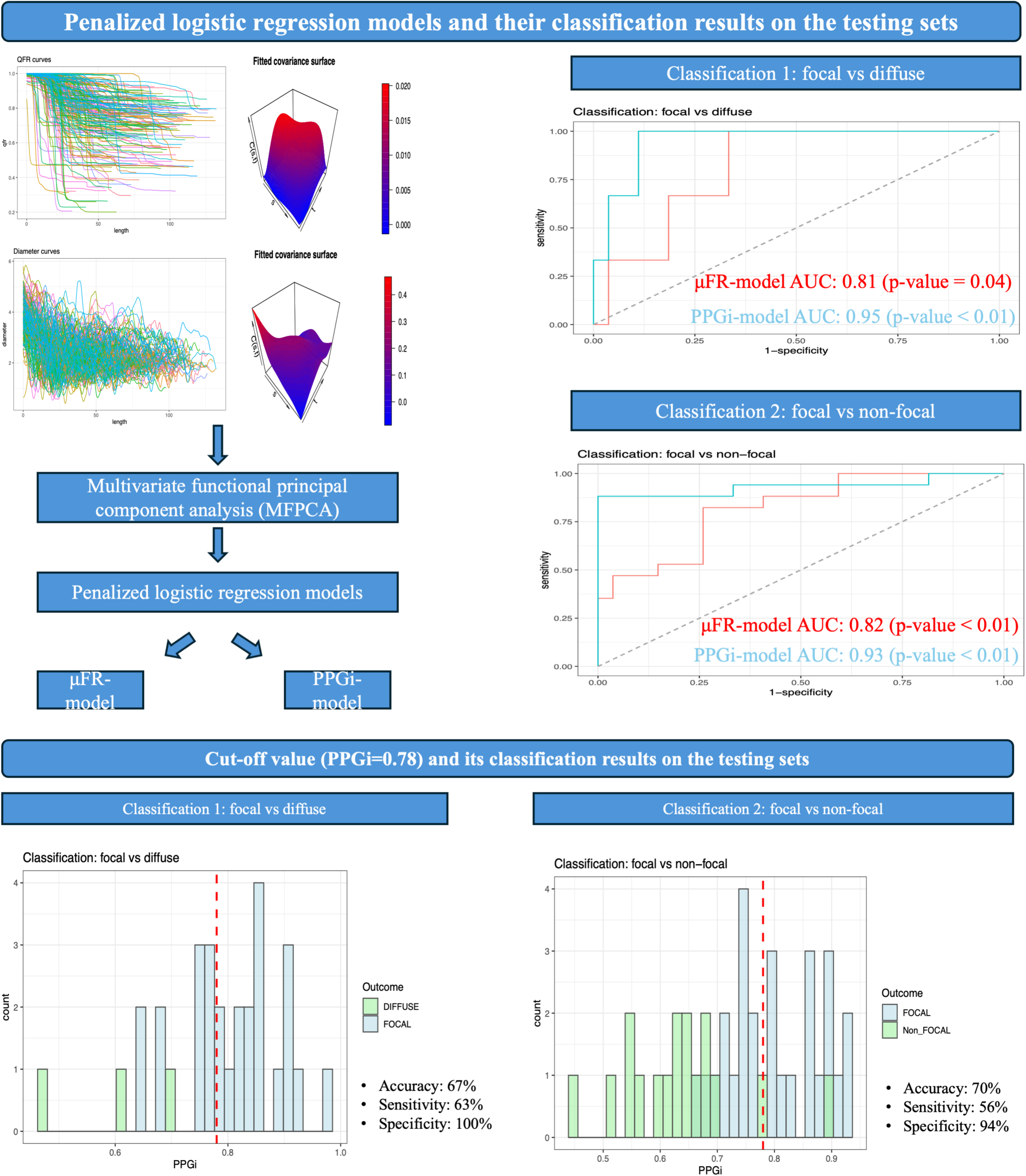
Central Illustration. Penalized logistic regression models and their classification results on the testing sets. The top panel illustrates the classification results from two penalized logistic regression models, which are μFR-model and PPGi-model, respectively. MFPCA is applied to μFR and QCA diameter curves to derive scores, which are subsequently used as features in the two models. The μFR and QCA diameter curves and their corresponding covariance surface are provided in the top left corner. For the classification results, the receiver operating characteristic AUC indicates the PPGi-model outperforms μFR-model. The bottom panel shows the classification results by using cut-off value (PPGi=0.78, the red dashed lines). The histograms illustrate the distribution of PPGi values. For classification 1, all bars to the left (right) of the red dashed line are classified as diffuse (focal). For classification 2, all bars to the left (right) of the red dashed line are classified as non-focal (focal). AUC, area under the curve; μFR, Murray’s law-based quantitative flow ratio; PPGi, pullback pressure gradient index; MFPCA, multivariate functional principal component analysis; QCA, quantitative coronary analysis.

**Figure 3.**
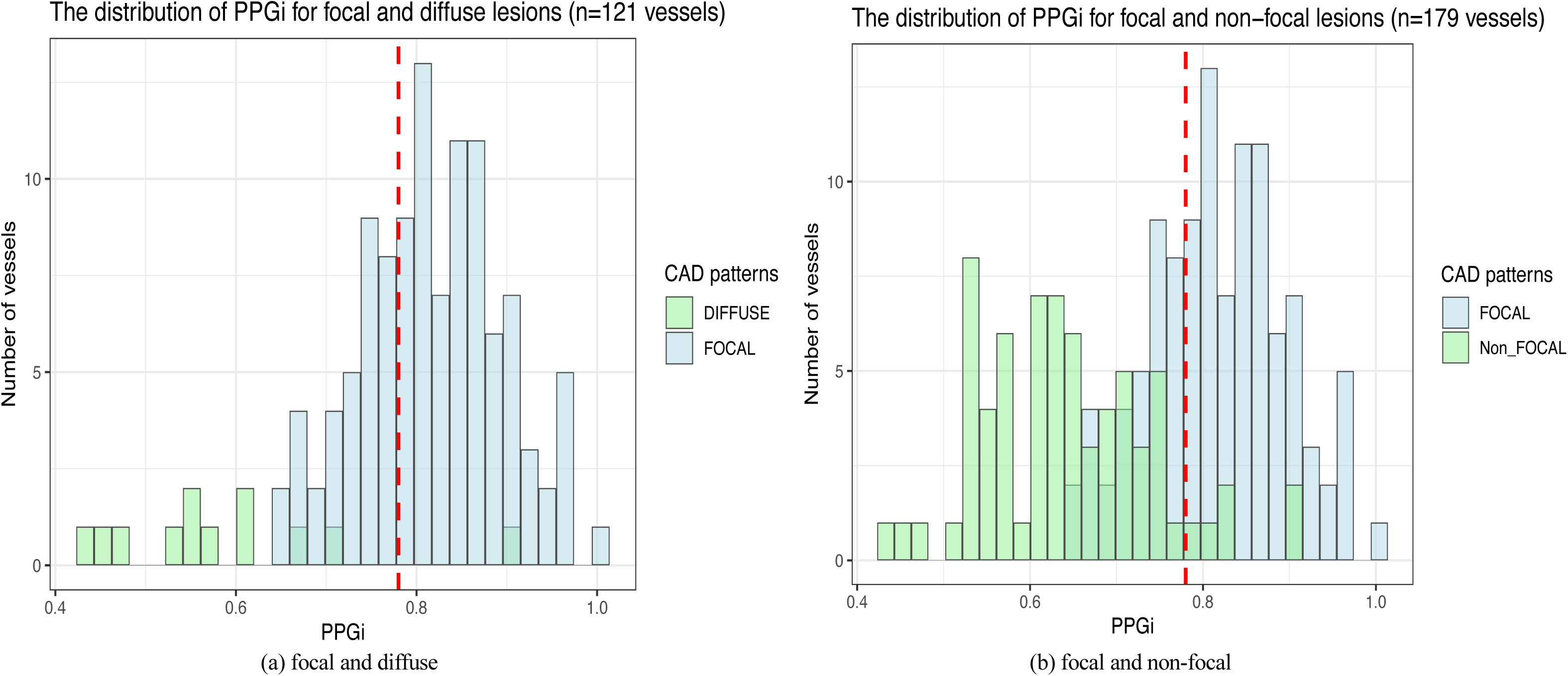
Distribution of PPGi and its cut-off value. The left panel shows the distribution of PPGi values for vessels with focal (blue) and diffuse disease (green). The right panel shows the distribution of PPGi values for vessels with focal (blue) and non-focal (green) disease. The red dashed lines represent the value of PPGi=0.78. CAD, coronary artery disease; PPGi, pullback pressure gradient index.

**Figure 4.**
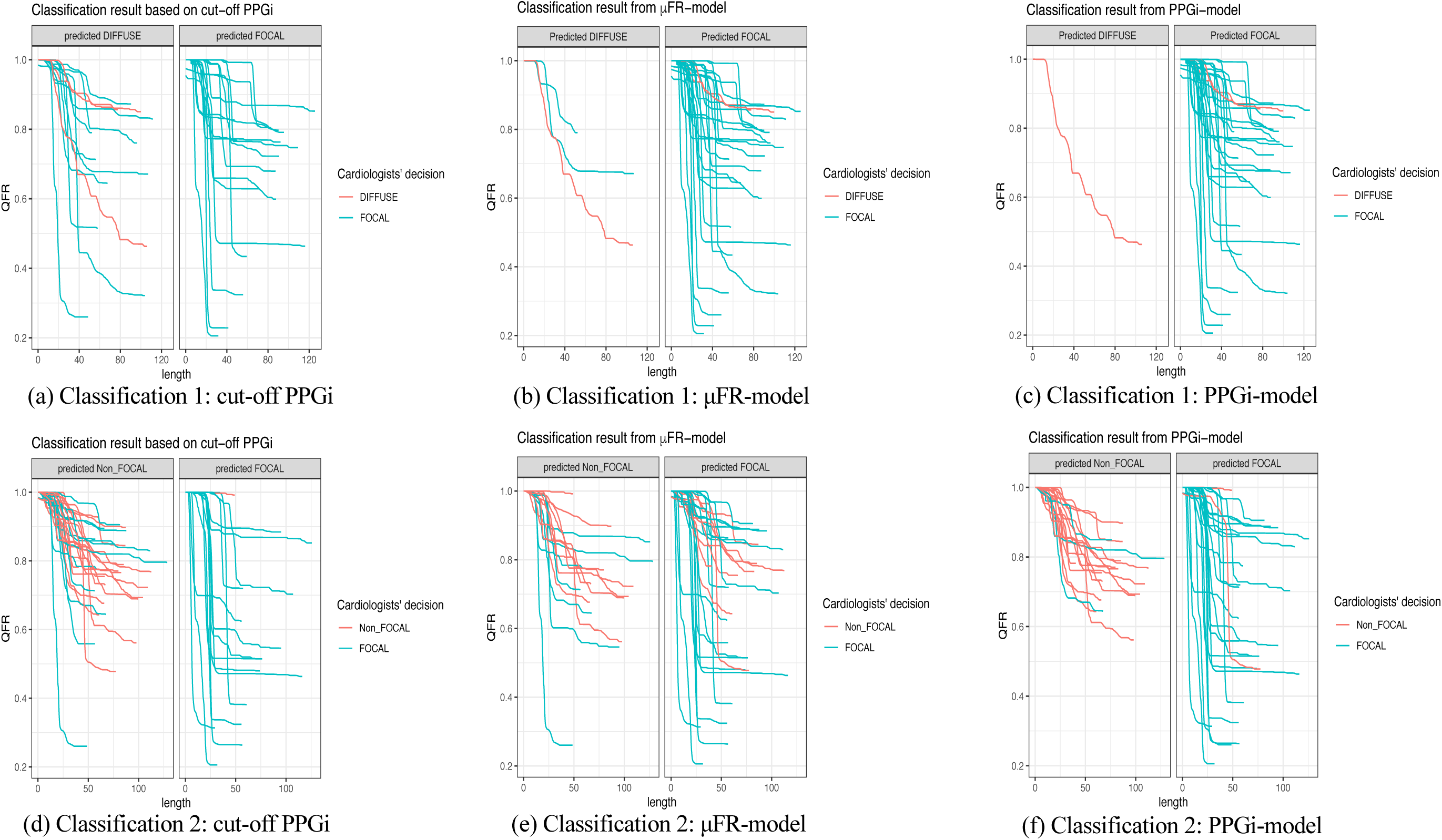
Classification results on the testing set based on the three methods. Classification 1: classification results on focal vs diffuse disease (for the testing set with 30 vessels). Red and blue lines represent the μFR values from vessels with diffuse and focal lesions, respectively, determined by the eight experienced interventional cardiologist. The predicted diffuse (focal) column shows the vessels that the classification method has predicted as having diffuse (focal) disease. Classification 2: classification results on focal vs non-focal disease (for the testing set with 44 vessels). Red and blue lines represent the μFR values from vessels with non-focal and focal lesions, respectively. The predicted non-focal (focal) column shows the vessels that the classification method has predicted as having non-focal (focal) disease. μFR, Murray’s law-based quantitative flow ratio; PPGi, pullback pressure gradient index; QFR, quantitative flow ratio.

### Classification using PPGi-model

The PPGi-model extended the μFR-model by including PPGi as one of the features. In order to compare the classification results, the same testing sets were used.

When considering the classification of focal vs diffuse, the PPGi-model successfully classified all focal lesions (sensitivity 100%), whereas only two diffuse were misclassified as focal (specificity 33%), see **Figure 4c**. PPV and NPV were 93% and 100%, respectively. The ROC curve with an AUC of 0.95 (p-value < 0.01) was provided in **Figure 2**. By including PPGi, the accuracy of PPGi-model improved to 93% (95% CI: 0.78 to 0.99), which outperformed μFR-model (97%, 95% CI: 0.70 to 0.96) and with the only PPGi (67%, 95% CI: 0.47 to 0.83).

In terms of the classification on focal vs non-focal, the PPGi-model resulted in an accuracy of 89% (95% CI: 0.75 to 0.96), with 24 focal (sensitivity 89%) and 15 non-focal (specificity 88%) correctly classified, see **Figure 4f**. Compared to the other two methods, the PPGi-model outperformed other models when dichotomizing CAD into focal and non-focal disease, with PPV of 92%, NPV of 83%, AUC of 0.93.

### Classification results after 500 iterations

Repeatedly splitting the data into training and testing sets helps to assess the robustness and generalizability of the model. **Table 2** lists the classification performance of the three methods after randomly splitting 121 vessels for focal and diffuse disease 500 times. Note that using the cut-off value of PPGi failed to provide the probability estimates, the AUC value was therefore not available. **Table 2** indicates that the PPGi-model performed the best classification, with an overall accuracy of 95%, PPV of 95% and NPV of 92%.

**Table 2.** Classification performance on focal vs diffuse (after 500 iterations)

|  | Accuracy | Sensitivity | Specificity | PPV | NPV | AC1 | AUC |
| --- | --- | --- | --- | --- | --- | --- | --- |
| PPGi=0.78 | 0.70 | 0.68 | 0.92 | 0.99 | 0.25 | 0.53 |  |
|  | (0.70-0.71) | (0.67-0.68) | (0.91-0.93) | (0.986-0.99) | (0.25-0.26) | (0.51-0.54) |  |
| μFR-model | 0.89 | 0.99 | 0.02 | 0.90 | 0.13 | 0.88 | 0.69 |
|  | (0.887-0.892) | (0.98-0.99) | (0.02-0.03) | (0.9-0.902) | (0.11-0.15) | (0.87-0.88) | (0.68-0.70) |
| PPGi-model | 0.95 | 0.99 | 0.51 | 0.95 | 0.92 | 0.94 | 0.92 |
|  | (0.94-0.95) | (0.992-0.995) | (0.49-0.54) | (0.947-0.952) | (0.91-0.94) | (0.93-0.94) | (0.90-0.93) |

AC, accuracy; AUC, area under the curve; $d\mu\text{FR}/ds$ , instantaneous $\mu\text{FR}$ gradient per unit length; $\mu\text{FR}$ , Murray's law-based quantitative flow ratio; NPV, negative predictive value; PPGi, pullback pressure gradient index; PPV, positive predictive value

Similarly, **Table 3** shows the classification performance of the three methods for focal vs non-focal cases after 500 iterations. In this case, the classification result derived from setting PPGi=0.78 as the cut off was better than μFR-model, whereas PPGi-model still outperformed the others with accuracy 84%, PPV 84% and NPV 84%.

**Table 3.** Classification performance on focal vs non-focal (after 500 iterations)

|  | Accuracy | Sensitivity | Specificity | PPV | NPV | AC1 | AUC |
| --- | --- | --- | --- | --- | --- | --- | --- |
| PPGi=0.78 | 0.77 | 0.68 | 0.91 | 0.93 | 0.64 | 0.54 |  |
|  | (0.76-0.77) | (0.67-0.68) | (0.90-0.92) | (0.92-0.93) | (0.64-0.65) | (0.53-0.55) |  |
| μFR-model | 0.68 | 0.81 | 0.47 | 0.71 | 0.62 | 0.41 | 0.73 |
|  | (0.67-0.68) | (0.80-0.82) | (0.46-0.48) | (0.706-0.714) | (0.61-0.63) | (0.40-0.42) | (0.72-0.73) |
| **PPGi-model** | 0.84 | 0.91 | 0.72 | 0.84 | 0.84 | 0.70 | 0.91 |
|  | (0.83-0.84) | (0.90-0.91) | (0.71-0.73) | (0.84-0.85) | (0.83-0.85) | (0.69-0.70) | (0.91-0.92) |

AC, accuracy; AUC, area under the curve; μFR, Murray's law-based quantitative flow ratio; NPV, negative predictive value; PPGi, pullback pressure gradient index; PPV, positive predictive value

## Discussion

In this study, we aimed to describe a novel model to classify CAD patterns based on machine-learning interpretation of angiography-derived physiology, and to preliminarily test its performance. This concept may translate into a precious tool to identify CAD patterns in a reproducible, generalizable and potentially automated way.

The main findings can be summarized as follows:

1. Pullback pressure gradient index with its validated cut-off (0.78) is a quantitative metrics that provides suboptimal accuracy in interpreting physiological pattern of CAD.
2. The penalized logistic regression models were developed to assess CAD patterns. Without including PPGi, the μFR-model demonstrated lower performance compared to PPGi.
3. Including PPGi to the penalized logistic regression model significantly improved the classification performance, leading to the best result among the three methods. The PPGi-model achieved an accuracy, PPV and NPV of 95% (95% CI: 0.94 to 0.95), 95% (95% CI: 0.947 to 0.952), 92% (95% CI: 0.91 to 0.94) for the classification of focal vs diffuse; and 84% (95% CI: 0.83 to 0.84), 84% (95% CI: 0.84 to 0.85), 84% (95% CI: 0.83 to 0.85) for the classification of focal vs non-focal patterns.

Functional patterns of CAD provide crucial information for PCI procedural planning and appropriateness. Diffuse disease poses several challenges in patients undergoing PCI including long coronary segments requiring treatment, lack of adequate landing zone for stenting and impairment of coronary flow related to the diffuse atherosclerotic burden along the vessel (22, 28, 29). Moreover, diffuse disease is associated with higher risk of suboptimal functional result and residual symptoms after PCI (21, 30, 31). Therefore, the determination of the distribution pattern of disease with the identification of focal lesions or segments of diffuse disease has become one of the cornerstones of decision making for myocardial revascularization.

Traditionally, at the PW-pullback, focal disease is defined as an abrupt pressure drop (delta FFR ≥0.05 or delta iFR ≥0.03) within a short vessel segment (≤20 mm), whilst serial/tandem lesions are defined as two or more focal stenoses separated by a non-diseased vessel segment >20 mm. Of note, the observed distribution pattern often combines features of focal and diffuse disease patterns (22, 28, 29). However, the qualitative interpretation of CAD patterns is hampered by a significant inter- and intra-operator variability, being potentially influenced by the expertise of physicians (32). In our original investigation, a complete agreement was achieved in only one third of vessels, while in more than 17% less than 5/8 concordant interpretations were achieved. Such difficulties in standardizing CAD patterns definitions and classification, together with technical and economic issues related to the need for a PW and hyperemic agent, hamper the wide-spread adoption of coronary physiology in real-world practice (33).

To overcome some of these limitations and standardize physiological pattern interpretation, quantitative metrics have been developed and validated. PPGi quantitatively measures the physiological distribution of coronary plaques along the vessel and is capable of distinguishing between focal and diffuse disease. Collet et al., in their preliminary experience, demonstrate PPGi reclassifies patterns interpretation in more than 30% of the lesions compared with angiography and it may facilitate the interpretation of FFR pullback traces (34). Another quantitative metric is the instantaneous FFR gradient per unit of time (dFFR[t]/dt), which allows identificationlocal disease severity in terms of major gradients that are associated with better PCI functional result (4, 6–11, 22). Both these metrics are calculated based on PW-pullback performed during continuous hyperaemia and require automatized motorized PW-pullback, further limiting the applicability in real-world practice.

Angiography-derived physiological indices, such as the μFR have been recently developed and validated to overcome several limitations of PW-assessment and lack of adoption in the real-world practice (3, 14, 15). μFR results in an inherently co-localized virtual pullback that allows to qualitatively interpret the physiological pattern of disease (focal vs. diffuse vs. mixed vs. serial) (2, 3, 16, 17).

The assessment of patterns based on angiography-derived physiology can be performed qualitatively by visual interpretation of the virtual pullback (35) or quantitatively with the μFR-PPGi. A cut-off value μFR-PPGi<0.78 was validated and seen to interact with long-term adverse events following PCI (21).

According to our analysis, PPGi derived from μFR virtual pullback provides suboptimal accuracy in interpreting physiological pattern of CAD, while our penalized logistic regression model was seen to provide improved classification performance, leading to the best result among the three methods, both when distinguishing between focal vs. diffuse and/or focal vs. non-focal disease. This model is conceived as a powerful tool allowing a simple, fast and reliable interpretation of CAD patterns, aiming to enhance precision medicine in terms of accurate diagnosis, procedural planning and tailored treatment. Importantly, the application of MFPCA and penalized logistic regression models present a potentially novel approach for CAD classification. The scores estimated from MFPCA summarized the joint variation between QFR and vessels’ diameter, and therefore, were included to the penalized logistic regression models to enhance the overall performance. Notably, the models can be reproduced and generalized across a range of study samples. In contrast, generalizing the cut off PPGi values to other study populations would be problematic, especially when they are derived from a specific study sample, such as the mean or median values (6, 7).

### Study limitations

Our study has limitations. The first limitation is the imbalanced number among four physiological CAD patterns. Due to the predominance of vessels with focal (61%) and mixed (23%) lesions over those with serial (9%) and diffuse (7%) disease, a multinomial logistic regression model would fail to provide reliable accuracy for the two minority classes. Therefore, it is more reasonable to fit a binomial logistic regression model by combining the mix, serial and diffuse disease as a non-focal class.

The assessment of CAD patterns from an individual expert cardiologist are likely to be subjective.

However, using an independent panel of eight experienced cardiologists, we found 83% of the vessels were labelled as having the same CAD pattern by a majority of experts (Figure 1). Furthermore, the remaining 32 vessels (yellow bars) without majority agreement underwent a second stage assessment to ensure the final labelled pattern was agreed upon. The lack of full agreement among the cardiologists, indicates that the decisions for these vessels were partly influenced by individual perspective. However, this fact highlights the requirement for an objective ML approach such as that developed in this study.

Finally, the cut-off values of PPGi, as they may be sensitive to the changes in population data. On the other hand, the machine learning method based on penalized logistic regression models should be considered a robust tool for the assessment of CAD patterns, and further studies would be required to validate its robustness across different study populations.

### Conclusions

Qualitative interpretation of physiological patterns of CAD is hampered by significant inter-observer variability. Angiography-derived PPGi, provides suboptimal accuracy in interpreting physiological pattern of CAD. Our study introduced a novel machine learning tool for CAD pattern classification using angiography-derived physiology. The penalized logistic regression models, particularly those incorporating PPGi, demonstrated superior performance in distinguishing focal from diffuse disease. This method offers a reliable, reproducible, and generalizable approach to enhance precision in myocardial revascularization, potentially addressing limitations in current qualitative assessments and improving patient outcomes.

### Clinical perspectives

This study proposes a novel machine learning-based method to classify physiological patterns of CAD using angiography-derived physiological indices, overcoming conventional metrics like FFR and angiography-derived PPGi. FFR and PPGi assessments have limitations, such as requiring dedicated pressure wires, hyperemic agents, and motorized pull-back, which restrict their use in clinical practice. The study introduces a penalized logistic regression model that incorporates PPGi and other vessel characteristics to improve the classification of CAD patterns into focal, diffuse, and non-focal types. The model was shown to outperform traditional PPGi-based classification, offering higher accuracy, sensitivity, and specificity. This machine learning approach has the potential to enhance decision-making in PCI by providing a more reliable and reproducible method for CAD pattern interpretation, integrating advanced computational methods into cardiovascular diagnostics to improve the precision and efficacy of treatment planning and optimization.

## Data Availability

All data produced in the present study are available upon reasonable request to the authors

## Abbreviations

AUC: area under the curve
CAD: coronary artery disease
dFFR[t]/dt: FFR gradient per unit time
DS: degree of stenosis
FFR: fractional flow reserve
iwFR: instantaneous wave-free ratio
MFPCA: multivariate functional principal component analysis
μFR: Murray’s law-based quantitative flow ratio
NPV: negative predictive value
PCI: percutaneous coronary intervention
PPGi: pullback pressure gradient index
PPV: positive predictive value
PW: pressure-wire
QCA: quantitative coronary analysis
QFR: quantitative flow ratio

