## Supplemental Table 1 for "Validation of Machine-Learning Angiography-Derived Physiological Pattern of Coronary Artery Disease"

**Supplementary Materials**

*2. Methods*

*2.4. Statistical analysis*

*Multivariate functional principal component analysis*

As μFR and diameter values were collected based on a point-by-point functional relationship with the length of the vessel, multivariate functional principal component analysis (MFPCA) was applied to these multivariate functional data, which consisted of μFR and diameter curves. The covariance surfaces shown in Central Illustration suggested the μFR curves exhibited most significant variation at the end of the vessels, whereas the diameter curves experienced predominant variation at the beginning of the vessels. Two important outcomes, multivariate functional principal components (MFPCs) and their scores, were calculated from MFPCA. Specifically, each element of the MFPCs captured the primary patterns of variation in the μFR and diameter curves about their means, respectively. The scores for each vessel were numerical values, which reflected the joint variation between μFR and diameter curves. In other words, the key features of μFR and diameter curves for an individual vessel were summarized together by these scores. Moreover, we selected 40 vessels based on their MFPC1 scores, including 20 vessels with higher scores (red lines) and 20 vessels with lower scores (blue lines), see Supplemental Figure 1. The vessels with higher MFPC1 scores were likely to exhibit more significant decreases in μFR values and have narrower diameters compared to those with lower MFPC1 scores. Consequently, the scores were carried forward as features of CAD patterns in penalized logistic regression models.

*Penalized logistic regression models*

Penalized logistic regression models were fitted to perform two classifications, namely focal vs diffuse and focal vs non-focal. Elastic net regularization was applied to impose a penalty to shrink coefficients of the less important features towards zero (i.e. performing feature selection), which helped to reduce the complexity of the model and to prevent the model from overfitting. Let $\lambda$ be the regularization parameter which controls the overall strength of the penalty, and $\alpha$ be mixing parameter which bridges the gap between lasso regression ($\alpha=1)$ and ridge regression ($\alpha=0)$. For each observation, $w_{i}$ and $l\left( y_{i},\beta_{0}+\beta^{T}x_{i} \right)$ represent the weight and negative log-likelihood. In theory, the elastic net solves the problem:

$$\min_{\beta_{0},\beta} \frac{1}{N}\sum_{i=1}^{N} w_{i}l\left( y_{i},\beta_{0}+\beta^{T}x_{i} \right)+\lambda\left[ \left( 1-\alpha\right)||\beta{||}_{2}^{2}/2+\alpha||\beta{||}_{1} \right].$$

In addition to two existing models in the paper, a third model μFR/PPGi(cut-off)-model was developed, see Supplemental Table 1. The μFR/PPGi(cut-off)-model treated PPGi as a qualitative variable, for instance, categorizing lesions as either focal (PPGi ≥ 0.78) or diffuse (PPGi<0.78). Supplemental Figure 2 shows the regularization path for coefficients from the three models as the regularization parameter $\lambda$ varies. Additionally, Supplemental Figure 3 illustrates the process of tuning parameters selection (based on the cross-validation method) in the training model to achieve optimal accuracy. After selecting the best tuning parameters, the coefficients can be extracted and used to do the predictions.

*3. Results*

*Classification using μFR/PPGi(cut-off)-model*

Firstly, μFR/PPGi(cut-off)-model was applied to the same testing set (n=30) to distinguish focal from diffuse patterns. However, including a binary PPGi variable failed to distinguish the diffuse disease, as all the 30 vessels were classified into focal class, see the left plot of Supplemental Figure 2. Nevertheless, μFR/PPGi(cut-off)-model still achieved excellent performance with an accuracy of 90% (95% CI: 0.73 to 0.98). The Gwet’s AC1 value of 0.89 suggested a high level of agreement between the results from the model and the decisions made by the eight cardiologists. It was because the focal lesions (n=27, 90%) were the most prevalent class in the testing set. In contrast, none of the diffuse lesions were identified, which caused zero specificity and undefined NPV.

On the other hand, μFR/PPGi(cut-off)-model utilized the same testing set (n=44) for focal and non-focal classification. The left plot of Supplemental Figure 2 shows only one non-focal patterns were misclassified as focal. The accuracy, sensitivity, specificity, PPV, NPV of the model were 70% (95% CI: 0.55 to 0.83), 56%, 94%, 94%, and 57% respectively.

### *Classification results after 500 iterations*

Supplemental Table 2 and Supplemental Table 3 list the classification performance of the four methods after running 500 iterations. There was no significant improvement for the classification on focal vs diffuse when including a qualitative PPGi variable to the μFR-model. In terms of the classification on focal vs non-focal, the μFR/PPGi(cut-off) model indeed improved the results compared to the μFR-model. However, the PPGi-model which included quantitative PPGi as one of the features, performed the best classification for the two cases.

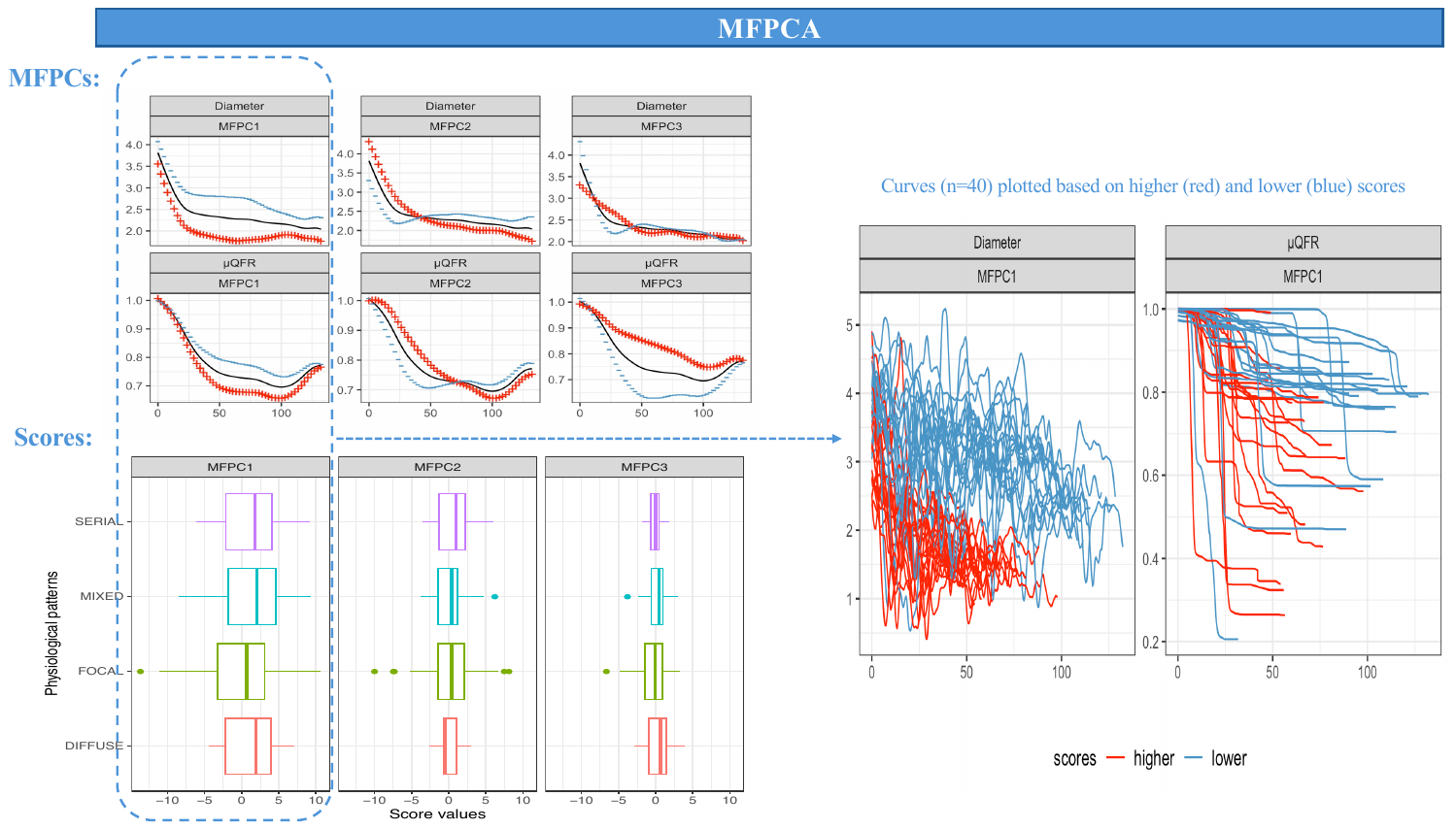
Supplemental Figure 1 Multivariate functional principal component analysis (MFPCA)

The first three MFPCs are provided in the top left corner, where the black lines represent the mean functions of diameter and QFR curves, and the red (blue) lines are the MFPCs added to (subtracted from) the mean functions. The boxplots for the corresponding scores for four CAD patterns are provided in the bottom left corner. 40 curves are randomly selected and plotted based on their first MFPC score, where the red (blue) lines represent the curve with higher (lower) score.

MFPCs, multivariate functional principal components; QFR, quantitative flow ratio; CAD, coronary artery disease.

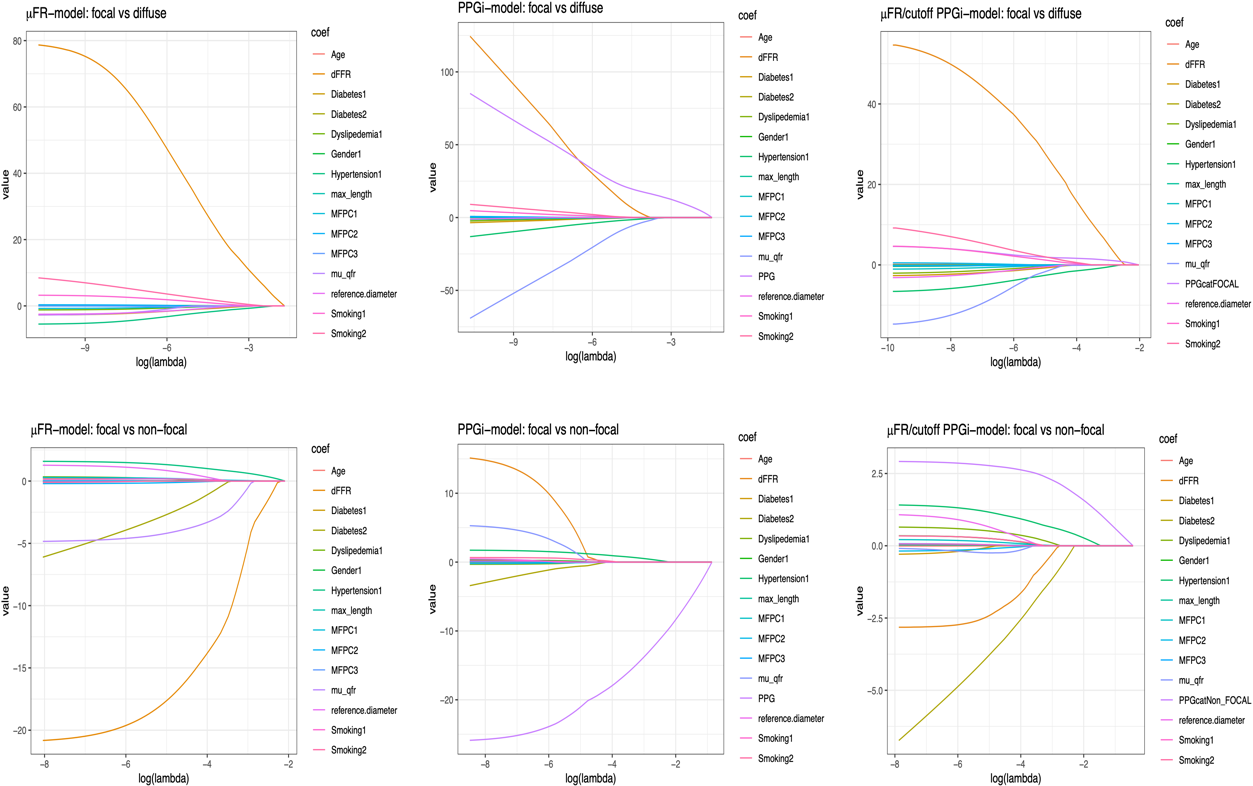

Supplemental Figure 2 Regularization paths for model coefficients

The top and bottom panels show the regularization paths for the three models used for classification on focal vs diffuse and focal vs non-focal, respectively. Each coloured line shows the regularization path of one feature in the model as the regularization parameter varies.

μFR, Murray’s law-based quantitative flow ratio; PPGi, pullback pressure gradient index.

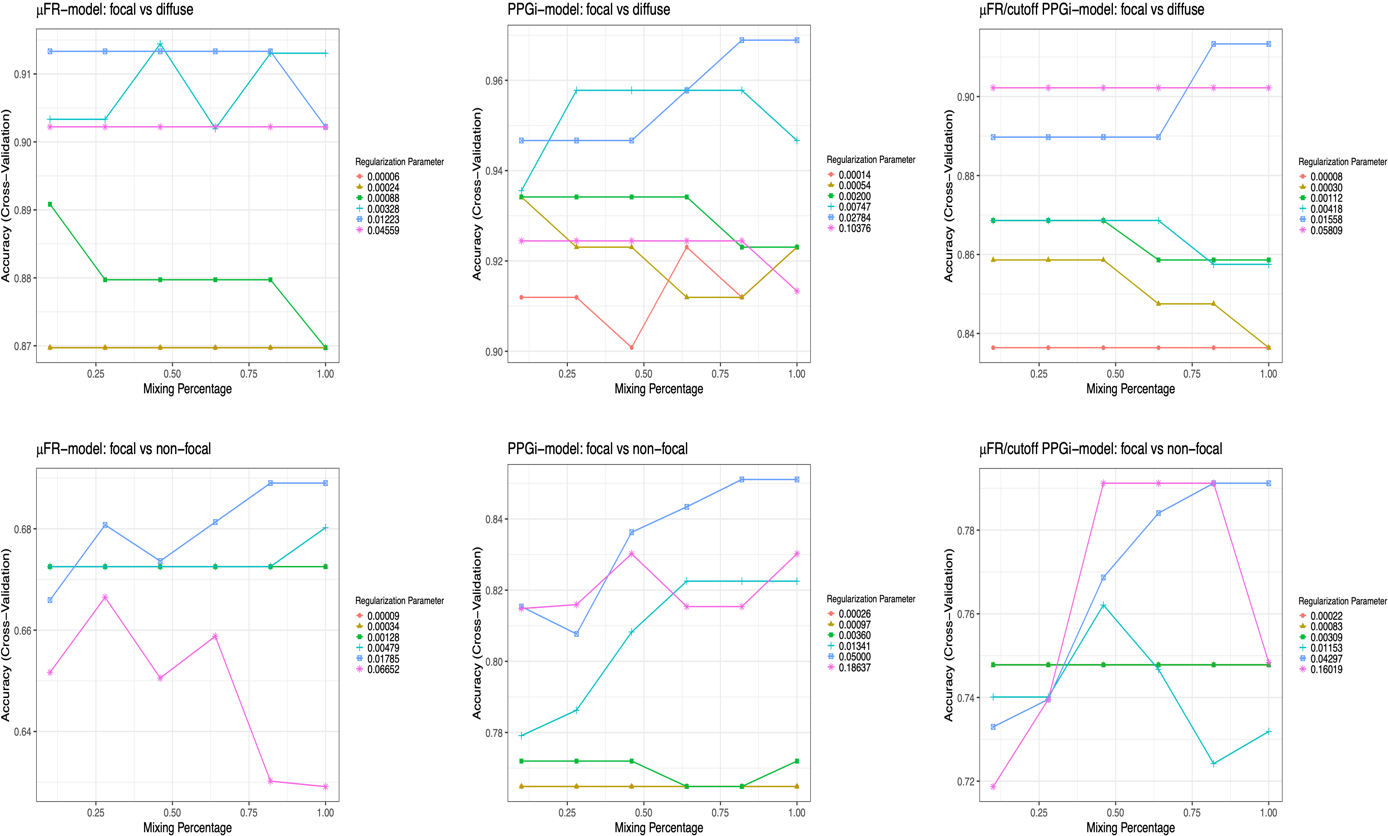

Supplemental Figure 3 The selection for tuning parameters

The top and bottom panels show the selection for tuning parameters in the three models used for classification on focal vs diffuse and focal vs non-focal, respectively. Each coloured line represents a different value of regularization parameter. The mixing percentage balances the lasso and ridge regression. The tuning parameters are selected based on highest accuracy.

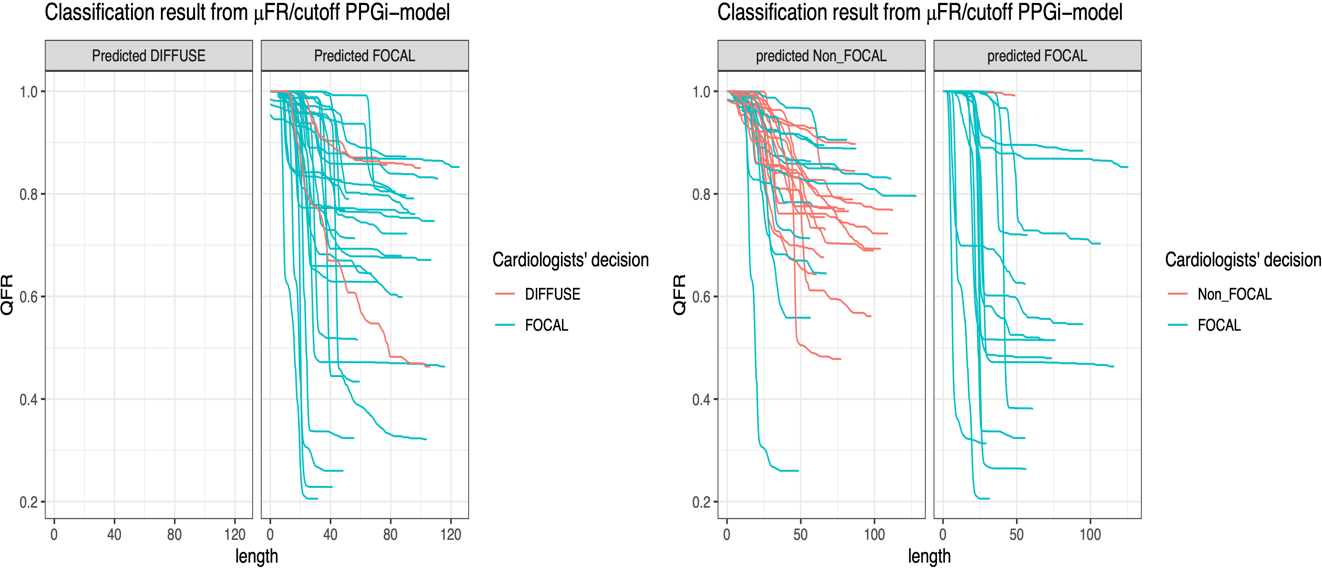

Supplemental Figure 4 Classification results for μFR/PPGi(cut-off)-model.

The red and blue lines in the left panel represent the QFR values from vessels with diffuse and focal lesions, respectively, determined by the eight experienced interventional cardiologist. The predicted focal (diffuse) column shows the vessels which are classified as focal (diffuse) by using the model. The red and blue lines in the right panel represent the QFR values from vessels with non-focal and focal lesions, respectively. The predicted focal (non-focal) column shows the vessels which are classified as focal (non-focal) by using the model.

Supplemental Table 1 Three penalized logistic regression models

| Features | | | | | | |
| --- | --- | --- | --- | --- | --- | --- |
|  | Vessels (2 variables) | Demographics  (6 variables) | MFPCA  scores  (3 variables) | Indices | | |
|  |  |  |  | dμFR /ds | μFR | PPGi |
| μFR-model | √ | √ | √ | √ | √ |  |
| PPGi-model | √ | √ | √ | √ | √ | Quantitative |
| μFR/PPGi(cut-off)-model | √ | √ | √ | √ | √ | Qualitative |

Supplemental Table 2 Classification performance on focal vs diffuse (after 500 iterations)

|  | Accuracy | Sensitivity | Specificity | PPV | NPV | AC1 | AUC |
| --- | --- | --- | --- | --- | --- | --- | --- |
| PPGi=0.78 | 0.70 | 0.68 | 0.92 | 0.99 | 0.25 | 0.53 |  |
|  | (0.697-0.710) | (0.672-0.687) | (0.910-0.934) | (0.986-0.990) | (0.246-0.257) | (0.513-0.540) |  |
| μFR-model | 0.89 | 0.99 | 0.02 | 0.90 | 0.13 | 0.88 | 0.69 |
|  | (0.887-0.892) | (0.983-0.989) | (0.015-0.031) | (0.900-0.902) | (0.108-0.145) | (0.871-0.878) | (0.682-0.703) |
| μFR/PPGi(cut-off) | 0.89 | 0.99 | 0.01 | 0.90 | 0.14 | 0.88 | 0.77 |
|  | (0.890-0.895) | (0.988-0.993) | (0.007-0.019) | (0.9-0.901) | (0.114-0.164) | (0.876-0.882) | (0.764-0.781) |
| PPGi-model | 0.95 | 0.99 | 0.51 | 0.95 | 0.92 | 0.94 | 0.92 |
|  | (0.943-0.948) | (0.992-0.995) | (0.490-0.538) | (0.947-0.952) | (0.907-0.940) | (0.934-0.940) | (0.904-0.926) |

Supplemental Table 3 Classification performance on focal vs non-focal (after 500 iterations)

|  | Accuracy | Sensitivity | Specificity | PPV | NPV | AC1 | AUC |
| --- | --- | --- | --- | --- | --- | --- | --- |
| PPGi=0.78 | 0.77 | 0.68 | 0.91 | 0.93 | 0.64 | 0.54 |  |
|  | (0.763-0.772) | (0.672-0.684) | (0.904-0.915) | (0.920-0.929) | (0.639-0.649) | (0.529-0.547) |  |
| μFR-model | 0.68 | 0.81 | 0.47 | 0.71 | 0.62 | 0.41 | 0.73 |
|  | (0.672-0.683) | (0.799-0.815) | (0.461-0.482) | (0.706-0.714) | (0.609-0.629) | (0.404-0.424) | (0.721-0.733) |
| μFR/PPGi(cut-off) | 0.75 | 0.74 | 0.75 | 0.84 | 0.66 | 0.51 | 0.81 |
|  | (0.740-0.749) | (0.730-0.748) | (0.741-0.767) | (0.832-0.845) | (0.649-0.662) | (0.498-0.516) | (0.808-0.818) |
| PPGi-model | 0.84 | 0.91 | 0.72 | 0.84 | 0.84 | 0.70 | 0.91 |
|  | (0.831-0.840) | (0.902-0.912) | (0.712-0.732) | (0.838-0.847) | (0.832-0.847) | (0.689-0.704) | (0.909-0.917) |
